# Clinical Characteristics of 2019 Novel Infected Coronavirus Pneumonia: A Systemic Review and Meta-analysis

**DOI:** 10.1101/2020.02.14.20021535

**Authors:** Kai Qian, Yi Deng, Yong-Hang Tai, Jun Peng, Hao Peng, Li-Hong Jiang

## Abstract

**Background:** A novel pneumonia associated with the 2019 coronavirus infected pneumonia (NCIP) suddenly broke out in Wuhan, China in December 2019. 37287 confirmed cases and 813 death case in China (Until 8th/Feb/2019) have been reported in just fortnight. Although this risky pneumonia with high infection rates and high mortality rates need to be resolved immediately, major gaps in our knowledge of clinical characters of it were still not be established. The aim of this study is to summaries and analysis the clinical characteristics of 2019-nCoV pneumonia.

**Methods:** Literatures have been systematically performed a search on PubMed, Embase, Web of Science, GreyNet International, and The Cochrane Library from inception up to February 8, 2020. The Newcastle-Ottawa Scale was used to assess quality, and publication bias was analyzed by Egger’s test. In the single-arm meta-analysis, A fix-effects model was used to obtain a pooled incidence rate. We conducted subgroup analysis according to geographic region and research scale.

**Results:** A total of nine studies including 356 patients were included in this study, the mean age was 52.4 years and 221 (62.1%) were male. The pooled incidences rate of symptoms as follows: pharyngalgia (12.2%, 95% CI: 0.087-0.167), diarrhea (9.2%, 95% CI: 0.062-0.133) and headache (8.9%, 95% CI: 0.063-0.125). Meanwhile, 5.7% (95% CI: 0.027-0.114) of patients were found without any symptoms although they were diagnosed by RT-PCR. In the terms of CT imaging examination, the most of patients showed bilateral mottling or ground-glass opacity, 8.6% (95% CI: 0.048-0.148) of patients with crazy-paving pattern, and 11.5% (95% CI: 0.064-0.197) of patients without obvious CT imaging presentations. The pooled incidence of mortality was 8.9% (95% CI: 0.062-0.126).

**Conclusions:** To our knowledge, this is the first evidence-based medicine research to further elaborate the clinical characteristics of NCIP, which is beneficial to the next step of prevention and treatment.

## INTRODUCTION

Since Dec 8, 2019, several cases of unknown pneumonia have been reported in Wuhan, Hubei province, China. On January 3, 2020, the 2019 novel coronavirus was identified in samples of bronchoalveolar lavage fluid from a patient in Wuhan and was confirmed as the cause of the NCIP^1^. This coronavirus (CoV) was named ‘‘2019 novel coronavirus’’ or ‘‘2019-nCoV’’ by the World Health Organization (WHO)^2^. Six kinds of human coronaviruses have been previously identified^3^. These include HCoV-NL63 and HCoV-229E, which belong to the Alphacoronavirus genus; and HCoV-OC43, HCoVHKU1, severe acute respiratory syndrome coronavirus (SARS-CoV), and Middle East respiratory syndrome coronavirus (MERS-CoV), which belong to the Betacoronavirus genus^4^ Furthermore, coronaviruses have become associated with deadly respiratory infections in humans following the emergence of SARS-CoV in Guangdong, China during 2002, which affected 8098 people in 37 countries^5^. There then followed the MERS-CoV outbreak^6^. Early in the 2019n-CoV outbreak, it has become clear that the virus can be transmitted from human-to-human^7^. A total of 28129 NCIP cases in the world have been confirmed, including 12 patients in the United States. According to the government report by the mayor of Wuhan, at a press conference on 26th January 2020, approximately five million residents left Wuhan for other provinces within China and thousands of people left Wuhan for other countries before the lockdown^7^. Therefore, all over the world will meet this public health issues probably.

Until 8th/Feb/2019, 76 literatures have been reported regarding the epidemiology and clinical features of pneumonia caused by 2019¬nCoV. After we reviewed all these literature research by the clinical studies, various clinical symptoms have been proposed as the typical features for the 2019nCoV, furthermore, some features are controversial in different clinic environments. Therefore, evidence-based medical clinical characters are required urgently. In this study, we did a systemic review and a single meta-analysis of the clinical features of 2019nCoV pneumonia.

## METHODS

This review was conducted according to Preferred Reporting Items for Systematic Reviews and Meta-Analyses (PRISMA) guidelines for systematic reviews and meta-analyses^8^. Keywords and study eligibility criteria were determined. The protocol for the review was registered with PROSPERO (Provisional registration number: 168532)

### Search Strategy

PubMed, Embase, Web of Science, GreyNet International (http://www.greynet.org/), and The Cochrane Library were searched for articles published until February 6 2020. Articles on 2019 novel coronavirus pneumonia, such as 2019nCoV, NCIP, and Wuhan pneumonia were also manually retrieved. To maximize search sensitivity, no filters or limits on language were applied (the retrieval process is shown in Figure 1).

**Figure 1.** PRISMA flow chart for article selection in the meta-analysis.

#### Selection Criteria

The inclusion criteria were as follows: (1) studies reporting information regarding NCIP; (2) those that were clinical studies or consecutive cases; (3) availability of clinical data can be drawn from the articles; (4) containing more than three cases. The exclusion criteria were as follows: (1) repeat articles, letters, editorials, and expert opinions; (2) studies without usable data; and (3) the articles published in languages other than English and Chinese.

### Data extraction

Two investigators (K.Q & Y.D) independently extracted data from eligible studies; disagreements were resolved by discussion with a third investigator. For each study, the following information was recorded: necessary information (e.g., first author, year of publication), research characteristics (e.g., RCT, case report, retrospective study, course of treatment), and study subject characteristics (e.g., gender, age, CT imagines, symptoms, therapies, and the incidence of complications).

### Quality control

The Newcastle-Ottawa Scale was used to assess quality^9^. Assessment scores of 0-3, 4-6, and 7-9 indicated poor, fair, and good studies, respectively. Discrepancies were resolved by consensus.

### Publication bias

Since approximate ten studies were included in each approach group, funnel plots were used to detect publication bias. Publication bias was analyzed using Egger’s linear regression test, which measures funnel plot asymmetry.

#### Statistical analysis

All statistical analyses were performed using Comprehensive Meta-analysis (Version 2). The results are expressed as incidences and 95% CIs. A random-effects model was used to perform the statistical analyses, and a chi-squared test and I^2^ statistic were used to assess the inter-study heterogeneity. I^2^>50% indicated that heterogeneity was not statistically significant^10^. In order to further explore the sources of heterogeneity and examine whether the results differed by study characteristics, subgroup analysis was performed according to geographic region (Wuhan area and outside Wuhan area), research scale (<50 cases and ≥50 cases)^10^.

## RESULTS

### Literature Search

We initially retrieved 145 articles of 2019 novel coronavirus pneumonia, 9 of which met the criteria for inclusion in our series. Reasons for exclusion included duplicate reports (n = 58), without clinical characteristics (n = 42), and other types of studies such as comments and letters to the editor (n = 36) (Figure 1).

### Study Characteristics

The methodological quality of the retained studies was high for observational studies (overall satisfactory quality of evidence; table 1). Among eligible literatures, three studies included individual patient data of 15 patients (table 2). Another six articles included 341 patients, however, there were no individual patient data. Among all of these studies, seven studies (90%) were from the Wuhan area; two studies which included one patient was from the outside of the Wuhan area. Study size ranged from 3 to 138 subjects, seven studies involving less than 50 cases and two studies involving more than 50 cases.

**Table 1.**
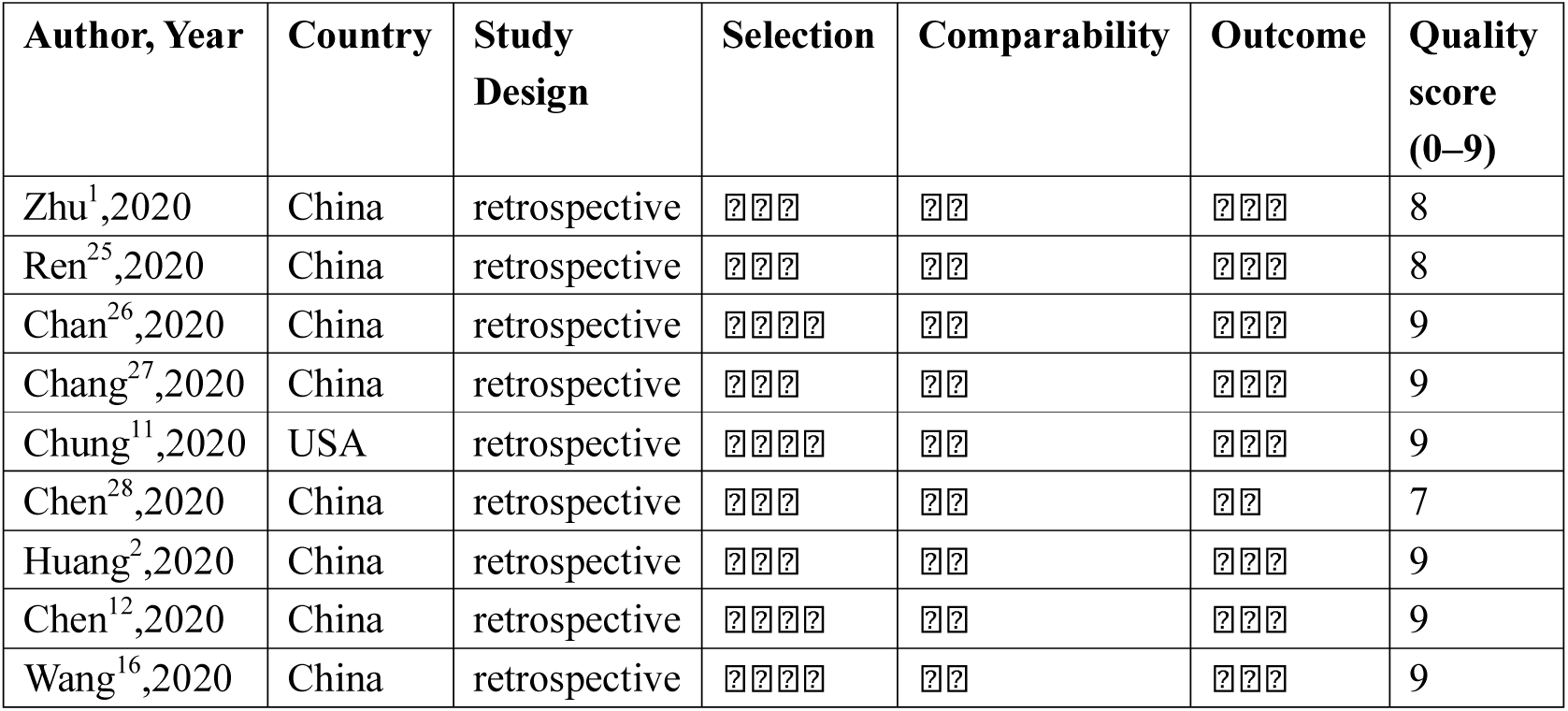
Characteristics and Quality assessment of included studies.

**Table 2.** Individual patient data of patients with NCIP.

| Author<br>s | Case<br>No | Age(<br>y)/se<br>x | Symptom | Onset to<br>treatmen<br>t interval | Highest<br>temperat<br>ure, °C | Underlying<br>conditions | WBC,<br>×10<sup>9</sup>/L | Number of<br>neutrophil<br>s, ×10<sup>9</sup>/L | Lymph<br>ocytes | CT<br>Imaging | Oxygen<br>therapy | ARDS | Outcome |
| --- | --- | --- | --- | --- | --- | --- | --- | --- | --- | --- | --- | --- | --- |
| Ren<sup>25</sup> | 1 | 65/M | C+E+A+F+H | 3 | 39.3 | Hypertension | 11.9 | 11.6 | 0.2 | BGGO | MV | Yes | Hospitalized |
| Ren<sup>25</sup> | 2 | 49/F | C+A+F+ | 5 | 38.5 | No | 8.3 | 7.6 | 0.44 | BGGO | NC | Yes | Hospitalized |
| Ren<sup>25</sup> | 3 | 52/F | C+A+F+M+H | 7 | 37.5 | No | 2.4 | 2.0 | 0.3 | BGGO | NC | No | Discharged |
| Ren<sup>25</sup> | 4 | 41/M | C+A+F+M | 6 | 39.0 | No | 6.6 | 5.0 | 0.98 | BGGO | NC | Yes | Hospitalized |
| Ren<sup>25</sup> | 5 | 61/M | C+E+A+F+M | 7 | N.A. | Tumor | 17.9 | 16.2 | 1.2 | BGGO | MV | Yes | Died |
| Chan<sup>26</sup> | 1 | 65/F | C+F+M | 5 | 39.0 | Hypertension<br>, Tumor | 4.8 | 4.0 | 0.6 | BGGO | N.A. | No | Hospitalized |
| Chan<sup>26</sup> | 2 | 66/M | C+F+M | 6 | 39.0 | Hypertension | 4.2 | 3.2 | 0.7 | BGGO | N.A. | No | Hospitalized |
| Chan<sup>26</sup> | 3 | 37/F | F+P+M+D | 4 | 36.2 | No | 5.6 | 3.1 | 2.6 | BGGO | N.A. | No | Hospitalized |
| Chan<sup>26</sup> | 4 | 36/M | C+F+D+P | 3 | 36.5 | Sinusitis | 11.4 | 8.1 | 2.7 | UGGO | N.A. | No | Hospitalized |
| Chan<sup>26</sup> | 5 | 10/M | Normal | N.A. | 36.5 | No | 6.5 | 3.2 | 2.8 | BGGO | N.A. | No | Hospitalized |
| Chan<sup>26</sup> | 6 | 7/F | Normal | N.A. | N.A. | No | Normal | Normal | N.A. | Normal | N.A. | No | Hospitalized |
| Chan<sup>26</sup> | 7 | 63/F | C+F+M | N.A. | 39.0 | Diabetes | 4.3 | 2.7 | 1.2 | BGGO | N.A. | No | Hospitalized |
| Zhu<sup>1</sup> | 1 | 49/F | C+F | 4 | N.A. | No | N.A. | N.A. | N.A. | BGGO | NC | No | Hospitalized |
| Zhu<sup>1</sup> | 2 | 61/M | C+F+A | 7 | N.A. | No | N.A. | N.A. | N.A. | BGGO | MV | Yes | Died |
| Zhu<sup>1</sup> | 3 | 32/M | C | N.A. | N.A. | No | N.A. | N.A. | N.A. | BGGO | NC | No | Hospitalized |

**Note:** C=Cough; E=Expectoration; A=Anhelation; F= Fever; M=Muscle pain/Fatigue; D=Diarrhea; H=Headache; P=Pharyngalgia; MV=Mechanical ventilation; ECMO=Extra-corporeal membrane oxygenation; NC=Nasal cannula; ARDS= Acute respiratory distress syndrome; BGGO=Bilateral mottling or ground-glass opacity; UGGO=Unilateral mottling or ground-glass opacity; CPP=Crazy-paving pattern

### Clinical symptom

There were 10 symptoms of NCIP which were reported. The pooled incidence rate was calculated for four symptoms: rhinorrhea (4.1%, 95% CI: 0.021-0.077), diarrhea (9.2%, 95% CI: 0.062-0.133), pharyngalgia (12.2%, 95% CI: 0.087-0.167), and headache (8.9%, 95% CI: 0.063-0.125). Interestingly, four patients were found without any symptoms although they were diagnosed by RT-PCR^11^. The pooled incidence rate of no obvious symptoms patients was 5.7% (95% CI: 0.027-0.114) for all studies. There was no significant heterogeneity (I^2^=47.67%, P=0.054) (Table 3). There was also no indication of publication bias as suggested by Egger’s test (P = 0.12). Among been reported clinical symptoms, evidence of heterogeneity was present in the symptoms of fever (I^2^ = 78.39%, P = 0.000), cough (I^2^ = 64.76%, P = 0.004) expectoration (I^2^ = 83.23%, P = 0.000) anhelation (I^2^ = 69.21%, P = 0.001), muscle pain (I^2^ = 74.9%, P = 0.000) and fatigue (I^2^ = 90.8%, P = 0.000). We found no identifiable sources of heterogeneity using subgroup analysis. The results of subgroup analyses according to geographic region and study scale are presented in Table 4.

**Table 3.** Descriptive Characteristics of Cases of Confirmed NCIP from inception up to February 7 2020 (n =356)

| Characteristic | Value | 95% CI | P value | I<sup>2</sup> |
| --- | --- | --- | --- | --- |
| **Age, year** |  |  |  |  |
| Mean | 52.4 | N.A. | N.A. | N.A. |
| Range | 6-92 | N.A. | N.A. | N.A. |
| **Sex, n (%)** | 356 | N.A. | N.A. | N.A. |
| Male | 221(62.1) | N.A. | N.A. | N.A. |
| Female | 135(37.9) | N.A. | N.A. | N.A. |
| **Clinical symptom, n (%)** |  | . |  |  |
| Fever | 313(86.2) | 0.683-0.948 | 0.000 | 78.39 |
| Rhinorrhea | 6(4.1) | 0.021-0.077 | 0.501 | 0.000 |
| Cough | 240(66.2) | 0.609-0.711 | 0.004 | 64.76 |
| Expectoration | 75(26.5) | 0.214-0.324 | 0.000 | 83.23 |
| Anhelation | 127(37.6) | 0.324-0.431 | 0.001 | 69.21 |
| Muscle pain | 79(27.5) | 0.225-0.331 | 0.000 | 74.90 |
| Fatigue | 132(47.1) | 0.406-0.538 | 0.000 | 90.80 |
| Diarrhea | 23(9.2) | 0.062-0.133 | 0.155 | 32.88 |
| Pharyngalgia | 30(12.2) | 0.087-0.167 | 0.051 | 48.20 |
| Headache | 29(8.9) | 0.063-0.125 | 0.672 | 0.000 |
| No obvious symptoms | 2(5.7) | 0.027-0.114 | 0.054 | 47.67 |
| **WBC, n (%)** |  |  |  |  |
| Increased | 71(20.7) | 0.167-0.253 | 0.466 | 0.000 |
| Decreased | 39(12.4) | 0.092-0.165 | 0.146 | 33.97 |
| **Neutrophils, n (%)** |  |  |  |  |
| Increased | 93(30.5) | 0.253-0.362 | 0.000 | 83.47 |
| **Lymphocytes, n (%)** |  |  |  |  |
| Decreased | 133(37.7) | 0.325-0.431 | 0.521 | 34.80 |
| **CT Imaging, n (%)** |  |  |  |  |
| BGGO | 307(74.8) | 0.681-0.804 | 0.002 | 66.88 |
| UGGO | 40(20.0) | 0.148-0.265 | 0.001 | 71.10 |
| CPP | 9(8.6) | 0.048-0.148 | 0.062 | 46.15 |
| Normal presentations | 9(11.5) | 0.064-0.197 | 0.002 | 67.16 |
| **Oxygen therapy, n (%)** |  |  |  |  |
| Nasal cannula | 268(74.4) | 0.696-0.788 | 0.688 | 0.000 |
| Mechanical ventilation | 78(22.8) | 0.186-0.275 | 0.591 | 0.000 |
| ECMO | 12(4.6) | 0.027-0.077 | 0.221 | 25.04 |
| **Clinical outcome, n (%)** |  |  |  |  |
| ARDS | 73(22.9) | 0.186-0.279 | 0.009 | 60.76 |
| Mortality | 26(8.9) | 0.062-0.126 | 0.432 | 0.176 |

**Note:** BGGO=Bilateral mottling or ground-glass opacity; UGGO=Unilateral mottling or
ground-glass opacity; CPP=Crazy-paving pattern; ECMO= Extra-Corporeal Membrane Oxygenation

**Table 4.** Subgroup analysis of incidence rate of clinical Characteristic.

|  | Geographic region |  |  |  |  |  | Study scale |  |  |  |  |  |
| --- | --- | --- | --- | --- | --- | --- | --- | --- | --- | --- | --- | --- |
| Characteristic | Wuhan area |  |  | Outside Wuhan area |  |  | < 50 cases |  |  | ≥50 cases |  |  |
|  | R%(95%CI) | I<sup>2</sup> | P | R%(95%CI) | I<sup>2</sup> | P | R%(95%CI) | I<sup>2</sup> | P | R%(95%CI) | I<sup>2</sup> | P |
| **Symptom** |  |  |  |  |  |  |  |  |  |  |  |  |
| Fever | 46.3(0.173-0.781) | 84.36 | 0.011 | 85.9(0.802-0.901) | 74.89 | 0.001 | 77.6(0.647-0.868) | 73.18 | 0.001 | 87.0(0.80-0.92) | 91.74 | 0.001 |
| Rhinorrhea | 10.4(0.026-0.336) | 0.000 | 0.644 | 3.2(0.02-0.07) | 0.000 | 0.652 | 5.5(0.022-0.132) | 0.000 | 0.691 | 3.1(0.012-0.075) | 62.42 | 0.103 |
| Cough | 50.0(0.293-0.707) | 0.000 | 0.640 | 67.3(0.62-0.72) | 70.11 | 0.003 | 63.9(0.545-0.727) | 36.47 | 0.150 | 67.4(0.609-0.733) | 92.23 | 0.000 |
| Expectoration | 8.8(0.018-0.345) | 0.000 | 0.403 | 27.3(0.22-0.33) | 88.56 | 0.000 | 36.4(0.262-0.480) | 77.24 | 0.000 | 21.9(0.164-0.285) | 93.64 | 0.000 |
| Anhelation | 49.4(0.267-0.724) | 70.84 | 0.064 | 37.0(0.32-0.43) | 72.20 | 0.001 | 52.9(0.427-0.628) | 53.73 | 0.044 | 31.2(0.256-0.374) | 0.000 | 0.980 |
| Muscle pain | 7.2(0.014-0.289) | 0.000 | 0.901 | 28.3(0.23-0.34) | 78.76 | 0.000 | 27.8(0.183-0.396) | 62.93 | 0.013 | 27.4(0.218-0.339) | 93.62 | 0.000 |
| Fatigue | 27.9(0.094-0.593) | 70.63 | 0.065 | 48.0(0.41-0.55) | 92.68 | 0.000 | 33.7(0.240-0.449) | 63.51 | 0.012 | 53.5(0.456-0.614) | 98.40 | 0.000 |
| Diarrhea | 17.8(0.057-0.437) | 27.52 | 0.240 | 8.4(0.056-0.125) | 34.46 | 0.173 | 11.2(0.059-0.204) | 6.75 | 0.376 | 8.2(0.051-0.131) | 79.60 | 0.027 |
| Pharyngalgia | 8.8(0.018-0.345) | 0.000 | 0.403 | 12.4(0.008-0.171) | 58.82 | 0.024 | 4.8(0.018-0.122) | 0.000 | 0.705 | 13.9(0.098-0.193) | 86.21 | 0.007 |
| Headache | 18.9(0.067-0.430) | 0.000 | 0.348 | 8.1(0.056-0.116) | 0.000 | 0.861 | 11.9(0.07-0.20) | 0.000 | 0.731 | 7.2(0.05-0.11) | 0.000 | 0.647 |
| No obvious symptoms | 18.0(0.050-0.475) | 51.00 | 0.153 | 3.4(0.014-0.081) | 31.36 | 0.189 | 8.8(0.040-0.181) | 14.76 | 0.317 | 0.4(0.001-0.03) | 0.00 | 0.869 |
| **WBC** |  |  |  |  |  |  |  |  |  |  |  |  |
| Increased | 10.4(0.026-0.336) | 0.000 | 0.644 | 21.1(0.17-0.26) | 4.19 | 0.394 | 23.3(0.164-0.320) | 0.000 | 0.680 | 19.4(0.148-0.250) | 66.8 | 0.083 |
| Decreased | 7.2(0.014-0.289) | 0.000 | 0.901 | 12.6(0.09-0.17) | 48.17 | 0.072 | 18.8(0.125-0.272) | 0.000 | 0.701 | 8.1(0.052-0.123) | 0.000 | 0.607 |
| Neutrophils |  |  |  |  |  |  |  |  |  |  |  |  |
| Increased | 10.4(0.026-0.336) | 0.000 | 0.644 | 31.4(0.26-0.37) | 86.65 | 0.000 | 41.4(0.302-0.535) | 78.32 | 0.000 | 26.7(0.211-0.331) | 93.62 | 0.000 |
| Lymphocytes |  |  |  |  |  |  |  |  |  |  |  |  |
| Decreased | 20.7(0.079-0.442) | 0.000 | 0.488 | 38.5(0.33-0.44) | 79.29 | 0.000 | 51.7(0.419-0.614) | 67.19 | 0.006 | 31.3(0.257-0.375) | 25.71 | 0.246 |
| **CT Imaging** |  |  |  |  |  |  |  |  |  |  |  |  |
| BGGO | 50.0(0.293-0.707) | 0.000 | 0.640 | 77.9(0.712-0.843) | 65.02 | 0.009 | 72.1(0.61-0.81) | 55.81 | 0.035 | 76.9(0.68-0.84) | 89.99 | 0.002 |
| UGGO | 39.5(0.131-0.739) | 87.69 | 0.004 | 19.0(0.138-0.255) | 66.29 | 0.007 | 14.7(0.084-0.245) | 61.83 | 0.015 | 23.1(0.161-0.320) | 89.99 | 0.002 |
| CPP | 10.4(0.026-0.336) | 0.000 | 0.644 | 8.2(0.043-0.150) | 58.74 | 0.024 | 11.6(0.064-0.201) | 0.000 | 0.640 | 0.4(0.001-0.029) | 0.000 | 0.869 |
| Normal presentations | 31.9(0.149-0.557) | 14.62 | 0.279 | 4.9(0.002-0.106) | 50.01 | 0.062 | 16.3(0.090-0.277) | 47.05 | 0.079 | 0.4(0.001-0.029) | 0.000 | 0.869 |
| **Oxygen therapy** |  |  |  |  |  |  |  |  |  |  |  |  |
| Nasal cannula | 91.2(0.655-0.982) | 0.000 | 0.403 | 73.9(0.689-0.784) | 0.000 | 0.840 | 70.5(0.613-0.783) | 0.000 | 0.642 | 76.4(0.705-0.813) | 0.000 | 0.851 |
| Mechanical ventilation | 8.8(0.018-0.345) | 0.000 | 0.403 | 23.2(0.190-0.281) | 0.000 | 0.662 | 26.6(0.192-0.357) | 0.000 | 0.701 | 20.8(0.161-0.265) | 20.91 | 0.261 |
| ECMO | 4.7(0.007-0.270) | 0.000 | 0.774 | 4.6(0.026-0.078) | 43.34 | 0.102 | 7.5(0.035-0.154) | 20.96 | 0.270 | 3.0(0.014-0.061) | 0.000 | 0.953 |
| **Clinical outcome** |  |  |  |  |  |  |  |  |  |  |  |  |
| ARDS | 4.7(0.007-0.270) | 0.000 | 0.774 | 23.5(0.19-0.29) | 65.10 | 0.009 | 32.8(0.238-0.434) | 53.79 | 0.043 | 18.6(0.141-0.242) | 0.00 | 0.640 |
| Mortality | 4.7(0.007-0.270) | 0.000 | 0.774 | 9.1(0.063-0.130) | 19.76 | 0.279 | 10.7(0.060-0.183) | 0.000 | 0.721 | 7.9(0.050-0.124) | 72.98 | 0.054 |

**Note:** BGGO=Bilateral mottling or ground-glass opacity; UGGO=Unilateral mottling or ground-glass opacity; CPP=Crazy-paving pattern; ECMO=Extra-Corporeal Membrane Oxygenation

### WBC

White blood cells were below the normal range in 39 patients (the pooled incidence rate was 12.4%, I^2^=33.97, P=0.146) and above the normal range in 71 patients (the pooled incidence rate was 20.7%, I^2^=0.000, P=0.466) (table 3). Lymphocytes were below the normal range in 133 patients, the pooled incidence of increasing neutrophils was 37.7% (95% CI: 0.325-0.431) (Table 3). 93 patients had neutrophils above the normal range, evidence of heterogeneity and publication bias was present in it (I^2^ = 83.47%, P = 0.01, egger’s test P=0.012). We found no identifiable sources of heterogeneity using subgroup analysis (Table 4).

### CT imaging

The CT imagines of NCIP patients were reported differently. By reviewing the literature, there are three common manifestations as follows: (1) bilateral mottling or ground-glass opacity, (2) unilateral mottling or ground-glass opacity, (3) crazy-paving pattern. Among these studies. 307 patients showed bilateral mottling or ground-glass opacity, 40 patients showed bilateral mottling or ground-glass opacity, and 9 patients with crazy-paving pattern (8.6%, 95% CI: 0.048-0.148). Additionally, Chen et. al. reported that pneumothorax occurred in one patient^12^. Evidence of heterogeneity was present in the bilateral mottling or ground-glass opacity (I^2^ = 66.88%, P = 0.002), and unilateral mottling or ground-glass opacity (I^2^ = 71.1%, P = 0.001). We found no identifiable sources of heterogeneity using subgroup analysis; Furthermore, there were nine patients with normal CT presentations during the period of NCIP. The pooled incidence rate of this normal CT imagint was 11.5% (95% CI: 0.064-0.197), significant heterogeneity was presented (I^2^ = 67.16%, P = 0.002). In subgroup analysis, heterogeneity was decreased, indicated that the heterogeneity may come from the geographic region (I2 = 14.62%, P = 0.279) and study scale (I2 = 47.05%, P = 0.079) (Table 4, Figure 2,3).

**Figure 2.** Forest plot of pooled to detect publication bias for normal CT presentation incidence rate. A. Subgroup analysis was performed according to geographic region (Wuhan area & outside Wuhan area). B. Subgroup analysis was performed according to research scale (<50 cases and ≥ 50 cases). The size of each square is proportional to the study’s weight. Horizontal lines indicate 95% CI. Diamonds indicate pooled incidence rate with its corresponding 95% CI. Egger test, P=0.21

**Figure 3.** Funnel plot to assess publication bias among studies. Each circle represents an identified study.

### Oxygen therapy

Nearly all the patients accepted oxygen therapy. Among these studies, there were 268 patients who used nasal cannula for 2-25 days, the pooled incidence was 74.4% (95% CI: 0.696-0.788) for all studies. 22.8% (95% CI: 0.186-0.275) patients used mechanical ventilation to assist ventilation, the inhaled oxygen concentration was 35-100%. Moreover, 12 patients were treated with extra-corporeal membrane oxygenation (ECMO), the pooled incidence was 4.6% (95% CI: 0.027-0.077). (Table 2). There was no evidence of heterogeneity and publication bias. The results of subgroup analysis according to geographic region and study scale are presented in Table 3.

### Clinical outcomes

Unfortunately, 26 died cases were reported, the pooled incidence of mortality was 78.1% (95% CI: 0.062-0.126), there was no significant heterogeneity (I^2^=39.64%, P=0.569). Since the course of treatment of NCIP is about three weeks until some articles published, some patients still accepted therapy in the hospital, the statistics on mortality may be inaccurate (Table 2). There was significant heterogeneity in the ARDS group (I^2^ = 60.76%, P = 0.01). We found no identifiable sources of heterogeneity using subgroup analysis (Table 3).

## DISCUSSION

In our research, the number of male patients more than female patients (62.1% vs 37.9%). This result is consistent with the gender distribution of MERS-CoV and SARS-CoV^13,14^. Meanwhile, Chen et.al also showed 2019-nCoV infection is more likely to affect males^12^. The reduced susceptibility of females to viral infections could be attributed to the protection from X chromosome and s sex-specific effects in infectious disease susceptibility^15^. On the contrary, a recent report that showed there was no difference in the proportion of men and women between ICU patients and non-ICU patients^16^. Although the mechanism of this difference cannot be explained at present, male patients should be paid more attention.

A recent study showed that nCoV was detected in stool samples of patients with abdominal symptoms ^17^. In our research, the pooled incidences rate of diarrhea was 7.8%, this results lower than the reported results of about 20-25% of patients with MERS-CoV or SARS-CoV infection^18^. Although the cause of this phenomenon is unclear, it suggests that we need to pay attention to patients with gastrointestinal symptoms and contact isolation should be taken. In addition, Four patients (5.7%) who with obvious symptoms were diagnosed by RT-PCR. Such patients will become a challenge in the future epidemic prevention process, which requires us to have detailed screening strategies, and we should be more vigilant in patients without obvious symptoms.

In terms of laboratory tests, the pooled incidence rate of lymphocytes reduce was 47.9%. On the other hand, the pooled incidence rate of increasing Neutrophils was 44.6%. These abnormalities are similar to those previously observed in patients with MERS-CoV and SARS-CoV infection^19^. These conclusions further confirm that lymphopenia along with neutrophilia was a feature of SARS-Cov, and 2019-nCoV might mainly act on lymphocytes, especially T lymphocytes^20^. Virus particles spread through the respiratory mucosa and infect other cells, induce a cytokine storm in the body, generate a series of immune responses, and cause changes in peripheral white blood cells and immune cells such as lymphocytes^12^. In addition, lymphopenia can be caused by glucocorticoids, and thus any debilitating condition has the potential to induce lymphopenia via stress mechanism involving the hypothalamic-pituitary-adrenal axis. Therefore, treatment with glucocorticoids complicated the issue regarding lymphopenia^21^.

Nine (11.5%) patients were diagnosed with NCIP, although the CT imaging was normal. This result reveals that the CT examination lacks complete sensitivity and cannot alone reliably fully exclude this disease, particularly early in the infection. Therefore, it is necessary to combine the CT examination with RT-PCR to make a definite diagnosis. Besides, Ground glass opacities, interlobular septal thickening, and consolidations were consistent HRCT manifestations in both metapneumovirus infection and SARS, the presence of crazy paving pattern is more suggestive of SARS^22^. Although the pooled incidence of crazy paving pattern (defined as thickened interlobular septa and intralobular lines with superimposed ground-glass opacification) was 8.4%, it is very important imagining for NCIP.

Until now, conclusions on the mortality of NCIP are inconsistent. The early two researches include 138 cases and 41 cases, the mortality was 4.3% and 15% respectively^2,16^. Nevertheless, compared with more than 10% mortality of SARS-CoV and 35% mortality of MERS-CoV^23^, 2019-nCoV has a lower case mortality. In our research, the pooled incidence mortality was 8.9% respectively. Although this conclusion is basically consistent with previous reports^2,12^, this result higher than the mortality reported by the Chinese government (2.44%). The reason for this phenomenon is attributed to two aspects. First of all, the single-arm meta-analysis is inherently less stable than the two-arm meta-analysis, but this is unavoidable due to no enough clinical data. Secondly, there were not enough diagnosis methods and treatment experiences about NCIP at the beginning of the outbreak. With the improvement of recognitions of NCIP and the clinical application of antiviral drugs such as Remdesivir^17^, the mortality will be further reduced.

A number of limitations need to be acknowledged. The limitations include those the number of included studies is small, thus limiting to the detection of the publication bias and leading to uncertainty of practical relevance of our meta-analysis. In addition, the clinical characteristics are related to many factors, such as basic physical condition, disease progress, examination and treatment conditions, etc. However, we were not able to conduct further subgroup analysis based on the abovementioned factor because most of the included studies did not separate the participants into different groups for outcome measurements. Third, significant heterogeneity remains a critical concern in this meta-analysis. To solve this problem, we used random-effects in meta-analysis and subgroup analysis was performed in this study^24^. Besides, we did not calculate the pooled incidence rate unless the source was identified by subgroup analysis. significant heterogeneity or a public basis^10^. Last but not least, the single-arm meta-analysis without a control group, causality is difficult to determine from the cases alone. However, all over the world, the onset of reactivation was relatively short and consistent. The strengths of this work include our ability to detect a serious question that was not observed during the clinical development program for NCIP.

In conclusion, the results of this single-arm meta-analysis and systemic review give us a quantitative pooled incidence rate of clinical characteristics of NCIP. It is clear that all these clinical characteristics has great potential to improve diagnosis and patient’s stratification in NCIP. It may also have a clinical impact as clinic features is routinely used in clinical practice, providing an unprecedented opportunity to improve decision support in NCIP for diagnosis or treatment at fast and low cost. The findings suggest that although most NICP patients have symptoms or abnormal CT imaging presentations, a few patients without any symptom or any abnormal CT imaging. Therefore, a prescriptive diagnosis process and the vigilance for NCIP is necessary. Besides, the digestive symptoms should be concerned, especially for the patients with the contact history of NICP. However, at the beginning of the epidemic, a lack of clinical research might influence the results of the meta-analysis. Further multivariate studies are warranted to corroborate the findings of this meta-analysis.

## Data Availability

The data used to support the findings of this study are available from the corresponding author upon request.

## Abbreviation List

ECMO: Extra-Corporeal Membrane Oxygenation
NCIP: 2019 coronavirus infected pneumonia
2019-nCoV: 2019 novel coronavirus
SARS-CoV: severe acute respiratory syndrome coronavirus
MERS-CoV: Middle East respiratory syndrome coronavirus
WHO: World Health Organization ()
ARDS: Acute respiratory distress syndrome

## ACKNOWLEDGEMENTS

We thank the patients, the nurses and clinical staff who are providing care for the patient, and all the people who fight with NCIP.

## REFERENCES

1. Zhu N, Zhang D, Wang W, et al. A Novel Coronavirus from Patients with Pneumonia in China, 2019. N Engl J Med. 2020.

2. Huang C, Wang Y, Li X, et al. Clinical features of patients infected with 2019 novel coronavirus in Wuhan, China. Lancet. 2020.

3. Wu A, Peng Y, Huang B, et al. Genome Composition and Divergence of the Novel Coronavirus (2019-nCoV) Originating in China. Cell Host Microbe. 2020.

4. Tang Q, Song Y, Shi M, Cheng Y, Zhang W, Xia XQ. Inferring the hosts of coronavirus using dual statistical models based on nucleotide composition. Sci Rep. 2015;5:17155.

5. Zhong NS, Zheng BJ, Li YM, et al. Epidemiology and cause of severe acute respiratory syndrome (SARS) in Guangdong, People’s Republic of China, in February, 2003. Lancet. 2003;362(9393):1353–1358.

6. Bawazir A, Al-Mazroo E, Jradi H, Ahmed A, Badri M. MERS-CoV infection: Mind the public knowledge gap. J Infect Public Health. 2018;11(1):89–93.

7. Khan S, Ali A, Siddique R, Nabi G. Novel coronavirus is putting the whole world on alert. J Hosp Infect. 2020.

8. Moher D, Liberati A, Tetzlaff J, Altman DG, Group P. Preferred reporting items for systematic reviews and meta-analyses: the PRISMA statement. J Clin Epidemiol. 2009;62(10):1006–1012.

9. Stang A, Jonas S, Poole C. Case study in major quotation errors: a critical commentary on the Newcastle-Ottawa scale. Eur J Epidemiol. 2018;33(11):1025–1031.

10. Higgins J, Thompson S, Deeks J, Altman D. Statistical heterogeneity in systematic reviews of clinical trials: a critical appraisal of guidelines and practice. J Health Serv Res Policy. 2002;7(1):51–61.

11. Chung M, Bernheim A, Mei X, et al. CT Imaging Features of 2019 Novel Coronavirus 1. (2019-nCoV). Radiology. 2020:200230.

12. Chen N, Zhou M, Dong X, et al. Epidemiological and clinical characteristics of 99 cases of 2019 novel coronavirus pneumonia in Wuhan, China: a descriptive study. Lancet. 2020.

13. Alqahtani FY, Aleanizy FS, Ali El Hadi Mohamed R, et al. Prevalence of comorbidities in cases of Middle East respiratory syndrome coronavirus: a retrospective study. Epidemiol Infect. 2018:1–5.

14. Leist SR, Cockrell AS. Genetically Engineering a Susceptible Mouse Model for MERS-CoV-Induced Acute Respiratory Distress Syndrome. Methods Mol Biol. 2020;2099:137–159.

15. Schurz H, Salie M, Tromp G, Hoal EG, Kinnear CJ, Moller M. The X chromosome and sex-specific effects in infectious disease susceptibility. Hum Genomics. 2019;13(1):2.

16. Wang D, Hu B, Hu C, et al. Clinical Characteristics of 138 Hospitalized Patients With 2019 Novel Coronavirus-Infected Pneumonia in Wuhan, China. JAMA. 2020.

17. Holshue ML, DeBolt C, Lindquist S, et al. First Case of 2019 Novel Coronavirus in the United States. N Engl J Med. 2020.

18. Assiri A, Al-Tawfiq JA, Al-Rabeeah AA, et al. Epidemiological, demographic, and clinical characteristics of 47 cases of Middle East respiratory syndrome coronavirus disease from Saudi Arabia: a descriptive study. Lancet Infect Dis. 2013;13(9):752–761.

19. Leist SR, Jensen KL, Baric RS, Sheahan TP. Increasing the translation of mouse models of MERS coronavirus pathogenesis through kinetic hematological analysis. PLoS One. 2019;14(7):e0220126.

20. Panesar NS. What caused lymphopenia in SARS and how reliable is the lymphokine status in glucocorticoid-treated patients? Med Hypotheses. 2008;71(2):298–301.

21. Roe MF, Bloxham DM, White DK, Ross-Russell RI, Tasker RT, O’Donnell DR. Lymphocyte apoptosis in acute respiratory syncytial virus bronchiolitis. Clin Exp Immunol. 2004;137(1):139–145.

22. Wong CK, Lai V, Wong YC. Comparison of initial high resolution computed tomography features in viral pneumonia between metapneumovirus infection and severe acute respiratory syndrome. Eur J Radiol. 2012;81(5):1083–1087.

23. Nkengasong J. China’s response to a novel coronavirus stands in stark contrast to the 2002 SARS outbreak response. Nat Med. 2020.

24. Imrey PB. Limitations of Meta-analyses of Studies With High Heterogeneity. JAMA Netw Open. 2020;3(1):e1919325.

25. Ren LL, Wang YM, Wu ZQ, et al. Identification of a novel coronavirus causing severe pneumonia in human: a descriptive study. Chin Med J (Engl). 2020.

26. Chan JF, Yuan S, Kok KH, et al. A familial cluster of pneumonia associated with the 2019 novel coronavirus indicating person-to-person transmission: a study of a family cluster. Lancet. 2020.

27. Chang, Lin M, Wei L, et al. Epidemiologic and Clinical Characteristics of Novel Coronavirus Infections Involving 13 Patients Outside Wuhan, China. JAMA. 2020.

28. Chen L, Liu HG, Liu W, et al. [Analysis of clinical features of 29 patients with 2019 novel coronavirus pneumonia]. Zhonghua Jie He He Hu Xi Za Zhi. 2020;43(0):E005.

